# UnitedMet harnesses RNA-metabolite covariation to impute metabolite levels in clinical samples

**DOI:** 10.1101/2024.05.24.24307903

**Authors:** Amy X. Xie, Wesley Tansey, Ed Reznik

## Abstract

Comprehensively studying metabolism requires the measurement of metabolite levels. However, in contrast to the broad availability of gene expression data, metabolites are rarely measured in large molecularly-defined cohorts of tissue samples. To address this basic barrier to metabolic discovery, we propose a Bayesian framework (“UnitedMet”) which leverages the empirical strength of RNA-metabolite covariation to impute otherwise unmeasured metabolite levels from widely available transcriptomic data. We demonstrate that UnitedMet is equally capable of imputing whole pool sizes as well as the outcomes of isotope tracing experiments. We apply UnitedMet to investigate the metabolic impact of driver mutations in kidney cancer, identifying a novel association between *BAP1* and a highly oxidative tumor phenotype. We similarly apply UnitedMet to determine that advanced kidney cancers upregulate oxidative phosphorylation relative to early-stage disease, that oxidative metabolism in kidney cancer is associated with inferior outcomes to combination therapy, and that kidney cancer metastases themselves demonstrate elevated oxidative phosphorylation relative to primary tumors. UnitedMet therefore enables the assessment of metabolic phenotypes in contexts where metabolite measurements were not taken or are otherwise infeasible, opening new avenues for the generation and evaluation of metabolite-centered hypotheses. UnitedMet is open source and publicly available (https://github.com/reznik-lab/UnitedMet).

## Introduction

Changes to metabolite pool sizes and metabolic flux are fundamental to numerous diseases and biological phenomena^1^, and by consequence measurement of metabolites themselves is critical to the discovery of disease biomarkers, therapeutic vulnerabilities, and mechanisms of action^2–4 5 6,7^. However, despite the translational value of metabolite measurement, large-scale profiling of metabolite levels in clinical specimens remains scarce due to the technical challenges associated with metabolomic measurements (e.g. the need for fresh, snap-frozen tissue, and the analytical challenges of measuring chemically diverse compounds) ^8^. Overcoming this data scarcity therefore comes with the potential reward of new access to the large space of underexplored, metabolite-centered biological hypotheses.

Two simultaneous and recent developments have now poised the metabolism field to overcome the lack of large-scale metabolite measurements. First, recent developments in machine learning have demonstrated the promise of using reference multimodal data (*i*.*e*. measurements of two or more distinct data modalities) to ultimately impute measurements of interest in single-modality data ^9,10^. For example, multi-modal learning methods for single cell multi-omics ^9–11^ have been successful at cross-modal prediction for single-modality datasets (e.g., protein prediction by jointly modeling with single cell RNAseq in TotalVI ^12^, single cell ATAC prediction via modeling with single cell RNA sequencing in MultiVI^13^). Second, we and other groups have identified both cancer-type-specific as well as lineage-agnostic patterns of RNA-metabolite covariation ^14–19^. Together, these developments suggest that suitably designed machine learning models may, by leveraging strong covariation between transcripts and metabolite pools, be able to predict otherwise unmeasured metabolite levels from matched single-modality transcriptomic data. Such a joint framework for modeling metabolic and RNA measurements would also produce a unified, low-rank representation of multimodal metabolite/RNA data, enabling downstream sample clustering, visualization and integration in a latent space.

Three key quantitative challenges must be addressed by multimodal models of metabolite/RNA levels. First, mass-spectrometry-derived metabolomics/isotope labeling data is predominantly reported in semi-quantitative relative abundances, impeding comparisons of identical metabolites/isotopologues across datasets (and of different metabolites within the same dataset). Second, different metabolomic measurement platforms often detect a subset of metabolites with limited overlap. As a result, each metabolic reference dataset exhibits a varying degree of missing measurements. Third, both metabolic and RNA modalities possess distinct sources of technical errors and noise which need to be suitably modeled.. Prior attempts at predicting metabolic profiles from RNA-seq data have had limited success, in part due to their inefficiency in addressing the aforementioned challenges. One method, reliant on correlation networks, struggled with missing values, resulting in a limited ability to predict cross-dataset outcomes for only 34 metabolites, with the highest Pearson’s ρ below 0.5 ^20^. Similarly, a different approach employing multivariate Lasso regression yielded poor performance, with a median *R*^2^ value of 0 for within-dataset prediction and an inability to perform cross-dataset prediction ^21^.

Here we present UnitedMet, a Bayesian probabilistic method for joint modeling of metabolic and RNA-seq data. UnitedMet addresses the above challenges by mapping both RNA and metabolite data onto a shared rank-transformed scale and inferring missing metabolic measurements in reference datasets. UnitedMet operates as a comprehensive framework at two levels. In the latent space, it learns a unified representation for both metabolic and RNA data, facilitating tasks like sample clustering and dataset integration. At a higher level, UnitedMet seizes on the strength of RNA-metabolite covariation to impute either metabolite pool sizes or isotopologue distributions from isotope labeling experiments directly from RNA abundance. We demonstrate that UnitedMet performs well on both imputation of pool sizes as well as imputation of isotope tracing experiments. We subsequently apply UnitedMet to identify the metabolite phenotypes of driver mutations in clinical specimens from patients with clear cell renal carcinoma (ccRCC), and study the metabolic phenotypes associated with metastatic disease in ccRCC.

## Results

### UnitedMet: A Bayesian probabilistic model for multimodal metabolic data analysis

UnitedMet is a Bayesian generative method that jointly models RNA-seq and metabolite data. The input to UnitedMet comprises the paired matrices of RNA counts (*X*) and total ion counts of metabolites/isotopologues (*Y*) from samples with both RNA-seq and metabolite data measured (defined as reference datasets) and single modality matrices with only RNA-seq data available (defined as target datasets) (**Figure 1A**). To map metabolite relative abundances and gene expression levels onto a shared measurement scale, we rank-transform the metabolite/isotopologue and gene expression levels across all the samples within each dataset. Such a rank transformation places the distribution of values for metabolite features onto a common, non-parametric scale which naturally accounts for the semi-quantitative nature of mass-spectrometry-based metabolomics data. UnitedMet then takes in an aggregate multiple-dataset matrix (*R*) containing the rankings data from both paired and single-modality samples. UnitedMet assumes observations are generated from a Plackett-Luce ranking distribution of a latent variable Z, which is the matrix product of a latent sample embedding matrix (*W*) and a latent feature embedding matrix (*H*) (**Figure 1B**). UnitedMet infers posterior distributions of gene expressions and metabolic profiles for all samples in the aggregate matrix and predicts metabolic profiles for single-modality samples using Stochastic Variational Inference (SVI). A hyperparameter λ, the number of latent embedding dimensions is selected by grid search. The output of UnitedMet is a fully-imputed multimodal data matrix, where any missing measurements from single modality data in the input matrix *R* are replaced with their posterior estimates.

**Figure 1.**
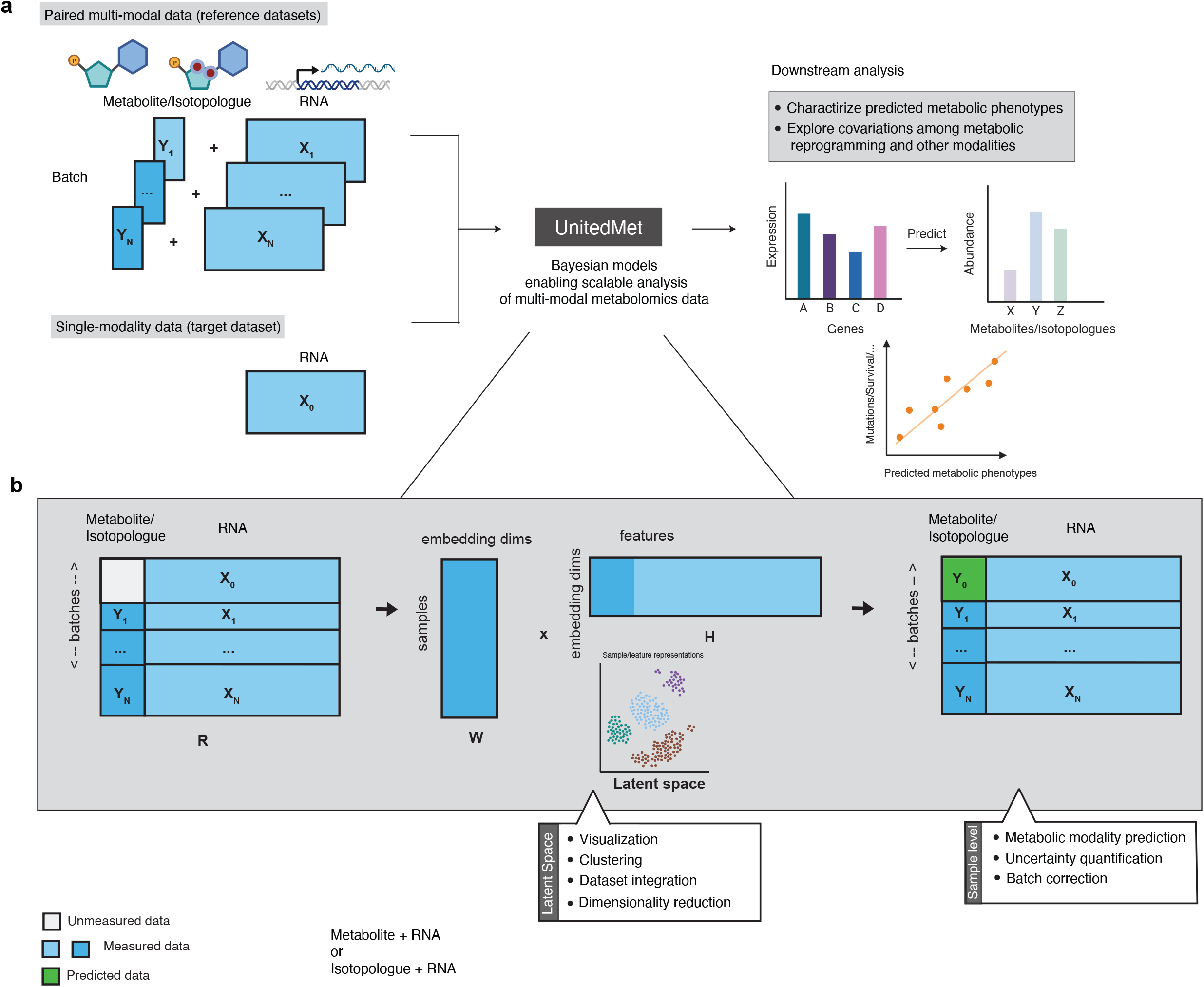
Overview of the UnitedMet method. **a)** Workflow of a metabolite imputation pipeline with UnitedMet. UnitedMet takes paired matrices of RNA counts (*X*) and total ion counts of metabolites/isotopologues (*Y*) (defined as reference datasets) and single modality matrices with only RNA-seq data available (*X*_0_) (defined as target datasets) as inputs. UnitedMet then normalizes and rank-transforms both RNA-seq and metabolic data. By probabilistic modeling, UnitedMet infers posterior distributions of metabolic profiles for single-modality target samples, which can be used in downstream analysis for biological hypothesis testing. **b)** Architecture of the UnitedMet model. An aggregate matrix (*R*) containing rankings data from both paired and single-modality samples is modeled with a Plackett-Luce ranking distribution based on latent variables derived from embedding matrices *W* and *H*. UnitedMet integrates transcriptomic and metabolic data into a common low-dimensional space for tasks like clustering and visualization. Next, UnitedMet imputes missing metabolite levels from gene expression data, offering predictions and uncertainty quantification.

UnitedMet provides a unified solution for multimodal metabolomic data analysis at two levels. First, UnitedMet learns a shared representation of both transcriptomic and metabolic data, including from samples where one type of measurement is missing, and integrates these data into a common low-dimensional latent space. Such a low-dimensional, integrated representation facilitates downstream tasks such as sample clustering and data visualization (**Figure 1B**). Second, by learning an unified representation of metabolomic and transcriptomic features from reference data, UnitedMet enables the imputation of otherwise unmeasured metabolite levels and/or isotopologue distributions from gene expression data alone, and delivers these predictions along with a quantification of their uncertainty. Together, these functions of UnitedMet enable the interrogation of metabolism, and the evaluation of hypotheses relying on metabolite measurements, in large, deeply-profiled cohorts of tumors otherwise lacking metabolomic data.

### UnitedMet accurately predicts metabolite levels from RNA sequencing data in human tumor samples

To evaluate UnitedMet’s capacity to predict metabolite abundances on real-world patient derived data, we first applied UnitedMet to four datasets of ccRCC patient samples with fully-paired RNA-seq and metabolomics profiles. The aggregated data contained two datasets from the NIH Clinical Proteomic Tumor Analysis Consortium (CPTAC) project, CPTAC (n=50, # metabolites = 183, # genes = 60483), CPTAC_val (n=71, # metabolites = 130, # genes = 60483), and two in-house datasets RC18 (n = 144, # metabolites=783, # genes = 22937), RC20 (n=76, # metabolites=1012, # genes = 22987). These data represented a typical use case for UnitedMet: while 20,171 genes were represented in all four datasets (corresponding largely to protein-coding genes uniformly measured across all data), only 86 (7% of the 1148 unique metabolites in the entire dataset) were measured in all four datasets.

We designed a benchmarking experiment to evaluate the performance of UnitedMet and comparator methods for the imputation of otherwise unmeasured metabolites. At each iteration of our benchmarking experiment, we treated three of the four ccRCC datasets as “reference” datasets for UnitedMet (in which both metabolomic and transcriptomic data is available), and treated the remaining ccRCC dataset as a “target” dataset (where only transcriptomic data was available). We subsequently trained four distinct UnitedMet models (one for each iteration of the benchmarking experiment, each with different hyperparameters λ) (**Figure S1A**) and evaluated the accuracy of UnitedMet metabolite predictions in the target dataset. For each metabolite, predicted levels from UnitedMet were compared with their ground-truth values by Spearman correlation (**Figure 2A**). We considered a metabolite well-predicted if the correlation between ground-truth and imputed abundance for that metabolite was positive and statistically significant (FDR-adjusted p-value < 0.05). UnitedMet successfully imputed between 48% and 67% of metabolites in the four target datasets (**Figure 2B**). We explored UnitedMet’s capacity to estimate model uncertainty by evaluating the standard error of 1000 draws from the posterior distribution of metabolite levels. We found that prediction uncertainty was negatively correlated with the prediction accuracy (**Figure S2B**), indicating that posterior uncertainty could guide the selection of reliable predictions for downstream analyses. Finally, we compared the performance of UnitedMet against two existing methods for prediction of metabolite abundance from gene expression with the same datasets: multivariable Lasso regression ^21^, and MIRTH ^22,23^ **(Methods)**. We used two metrics to quantify how well each method predicted metabolite abundance: the spearman rho among all predicted metabolites, and the number of well-predicted metabolites. UnitedMet outperformed the other methods in all 4 cross-validation datasets by both metrics (**Figure 2C, Figure S1B**).

**Figure 2.**
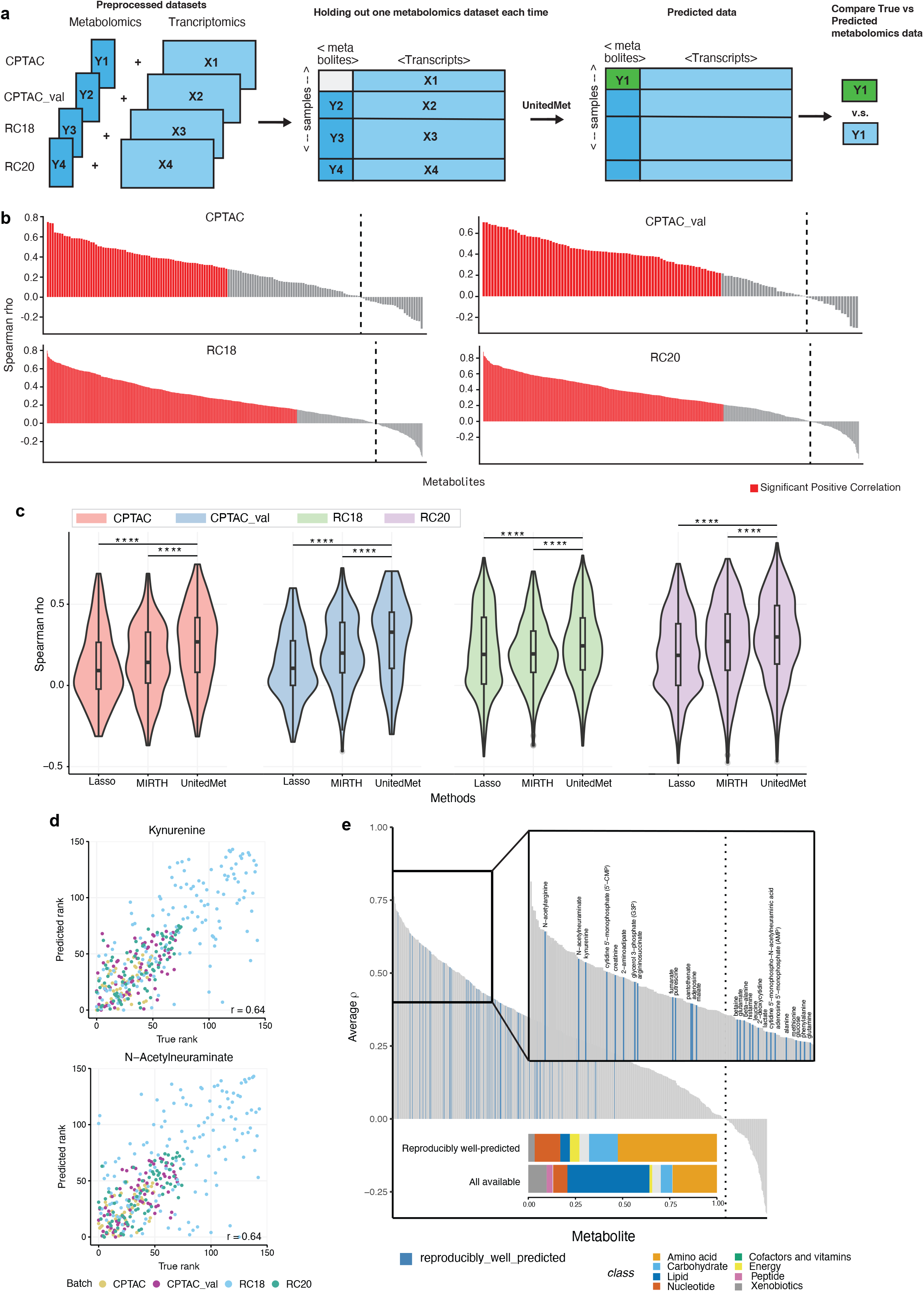
UnitedMet achieves high accuracy predicting metabolite levels in human tumor samples. **a)** Schematic of the benchmarking experiment to evaluate model performance in a cross-validation scenario. Each time, three out of four ccRCC datasets were designated as “reference” datasets, while the fourth dataset served as the “target” dataset with only transcriptomic data. The accuracy of UnitedMet’s predictions was then evaluated by comparing predicted metabolite abundances with their ground-truth levels. *X*: RNA-seq data; *Y*: Metabolomics data. **b)** The imputation performance for each dataset is assessed by spearman rho values between predicted values and their ground-truths across all simulated missing features. Metabolites with predicted ranks that show significant positive correlation (FDR-adjusted p < 0.05 and spearman rho > 0) with the actual ranks are labeled red. **c)** Performance of UnitedMet, multivariate Lasso regression, and MIRTH based on the metric: the spearman rho among all predicted metabolites. Significant difference was assessed by the Wilcoxon signed-rank test. **** denotes *P* < 0.0001. **d)** Correlation between actual and predicted metabolite ranks for two reproducibly well-predicted metabolites: kynurenine (top) and N-acetylneuraminate (bottom). Each point represents one sample in which the metabolite was measured and predicted. **e)** The imputation performance for each metabolite is summarized across datasets, with average spearman rho values plotted. A subset of consistently well-imputed metabolites is labeled, and those that are reproducibly well-predicted are marked in blue.

We next investigated the consistency of prediction accuracy across four datasets in UnitedMet. For each pair of two datasets from the four, we observed strong correlation between their metabolite-level prediction performances (Spearman correlation 0.32 - 0.83 in all pairwise comparisons, p < 0.001 in all pairwise comparisons; **Figure S2A**), confirming that well-predicted metabolites were highly consistent across all four datasets. For instance, kynurenine (average Spearman correlation 0.64, FDR-adjusted p-value < 0.05 in all 4 datasets) and N-acetylneuraminate (average spearman ρ = 0.64, FDR-adjusted p-value < 0.05 in all 4 datasets) exhibited robust prediction results across 4 datasets (**Figure 2C**). Combining these prediction results in four datasets, we labeled 59 metabolites as “reproducibly” well-predicted metabolites, indicating that they were well-predicted in at least 3/4 target datasets (**Figure 2D**). Reproducibly well-predicted metabolites were enriched for amino acids and carbohydrates, but depleted of lipids, relative to the full panel of metabolites (**Figure 2D)**.

As targeted mass spectrometry can only measure a specific class of metabolites, a related challenge is imputing a large panel of metabolites from a subset of measured metabolites and RNA-seq data. To address this, we extended UnitedMet’s capabilities by introducing a weighted loss function to address the imbalanced metabolomics and RNA-seq modalities. To benchmark the imputation accuracy, we randomly selected 50% of all measured metabolites as simulated-missing in each dataset. Once again, we found UnitedMet was the top performer on all datasets in terms of the same metrics mentioned above (**Figure S1C, Figure S1D**).

### UnitedMet can predict isotopologue distributions from RNA-seq data in vitro and in vivo

Unlike measurements of metabolite pool sizes, isotopologue distributions produced from steady-state isotopic labeling experiments capture the flow of nutrients through cellular metabolism. However, labeling experiments are technically challenging, and consequently there is even less publicly available isotopic labeling data (both in cell lines as well as in tissue specimens) than there is conventional metabolomic data. Motivated by the ability of UnitedMet to predict metabolite levels by jointly modeling metabolomics and RNA-seq data and the generalizability of our model, we hypothesized that UnitedMet might be able to predict isotopologue distributions from RNA-seq data. To test this hypothesis, we obtained three datasets with paired RNA-seq data and isotopic labeling data (measured by mass spectrometry). Dataset RCC contained renal cell carcinoma (RCC) tumor samples obtained from 76 patients receiving infusions of [U-^13^C]glucose prior to surgery^24^. A total of 64 isotopologues and 12300 genes were measured in the RCC dataset^24^. The other two datasets were composed of human non-small cell lung cancer (NSCLC) cell lines labeled by either [U-^13^C]glucose or [U-^13^C]glutamine: NSCLC-G (n=85), NSCLC-Q (n=85) ^3^. Both NSCLC datasets measured a total of 78 isotopologues and 16383 genes.

To evaluate UnitedMet’s performance of predicting isotopologue distributions, we conducted a simulation where 50% of the samples in a given dataset were randomly selected and treated as target data for UnitedMet (*i*.*e*. with isotopologue measurements masked, **Figure 3A**). The remaining 50% of samples were treated as a reference dataset for UnitedMet. We trained three distinct UnitedMet models (one for each dataset) with different hyperparameters λ (**Figure S3A**). UnitedMet was able to successfully impute 52% (RCC), 56% (NSCLC-G) and 63% (NSCLC-Q) of the held-out isotopologues (spearman ρ > 0, FDR-adjusted p-value < 0.05) respectively (**Figure 3B**). Citrate m+2, which reflects the contribution of glucose-derived carbon to the TCA cycle via pyruvate in [U-^13^C]glucose labeled data, was reproducibly predicted with high accuracy in both the in-vitro NSCLC dataset (spearman ρ = 0.44, p = 0.003) and the in-vivo RCC dataset (spearman ρ = 0.39, p = 0.01) (**Figure 3C**). In contrast, gene expression scores of either oxidative phosphorylation signature or TCA cycle signature, calculated directly from RNA-seq data, were not correlated to citrate m+2 labeling in these datasets (OXPHOS signature: spearman ρ = 0.16, p = 0.3 (NSCLC), spearman ρ = 0.22, p = 0.19 (RCC), **Figure 3C**; TCA cycle signature: ρ = 0.06, p = 0.3 (NSCLC), ρ = 0.25, p = 0.13 (RCC), **Figure S3C**). Similarly, lactate m+3, which reflects glucose contribution to glycolysis in [U-^13^C]glucose labeled data, was accurately predicted in the RCC dataset (spearman ρ = 0.43, p = 0.007), while a glycolysis gene expression signature was not correlated to lactate m+3 (spearman ρ = 0.05, p = 0.8; **Figure S3D**). Together, these results demonstrated UnitedMet can accurately predict isotopologues that characterize specific metabolic phenotypes, an achievement not possible with standard GSEA analysis of RNA-seq data.

**Figure 3.**
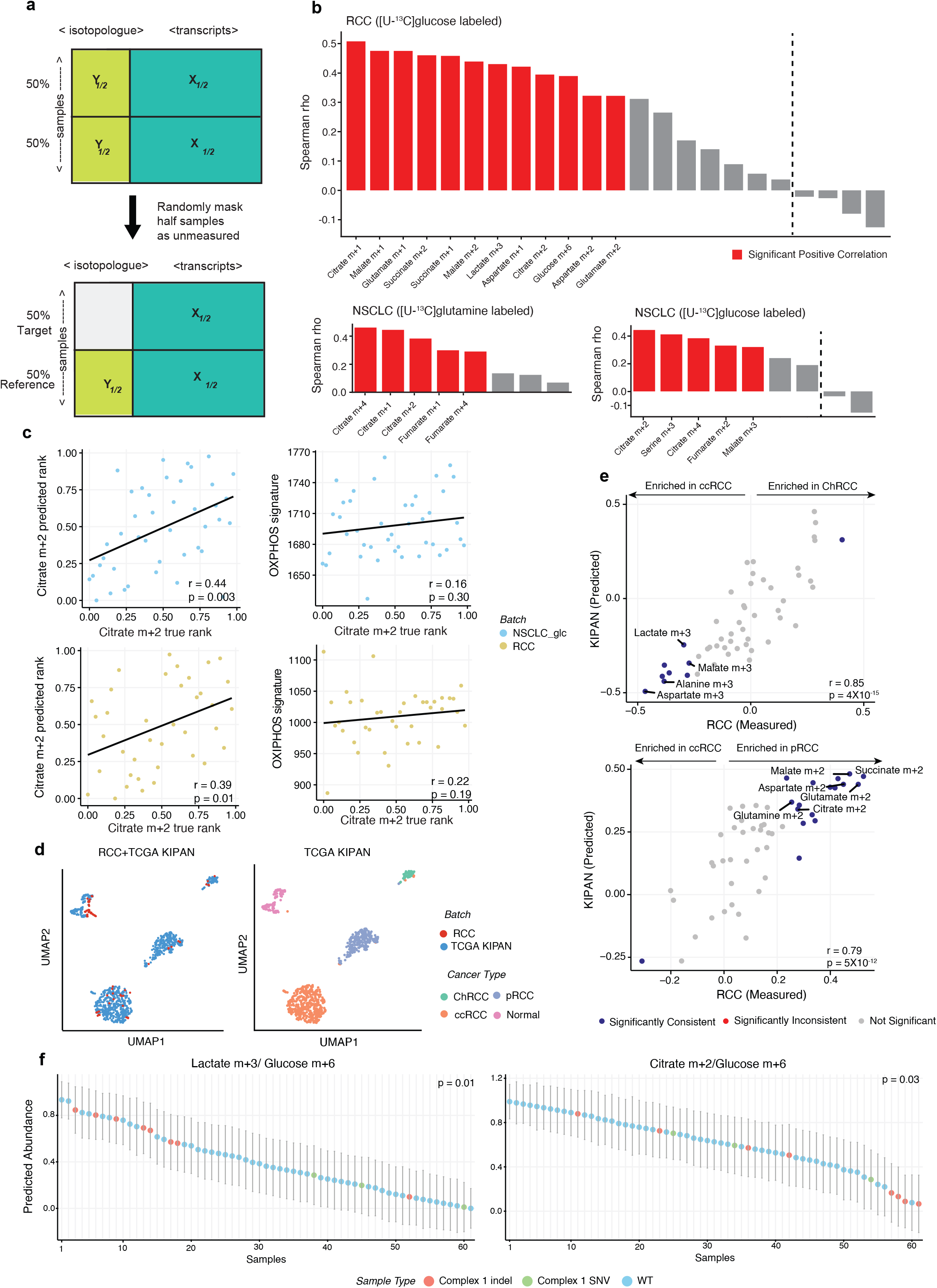
UnitedMet accurately predicts isotopologue distributions from RNA-seq data. **a)** Schematic of the benchmarking experiment to assess model performance on isotopologue predictions. 50% of the samples in a given dataset were randomly selected and treated as target data for UnitedMet (*i*.*e*. simulated as unmeasured). The remaining 50% of samples were treated as a reference dataset for UnitedMet. *X*: RNA-seq data; *Y*: Isotopologue data. **b)** Imputation performance for each dataset is evaluated using Spearman’s rho values between predicted values and their ground truths across all simulated missing features. Isotopologues with predicted ranks that exhibit a significant positive correlation with the actual ranks are marked in red. **c)** True ranks of citrate m+2 were well-predicted by UnitedMet but not by gene expression signature of Hallmark oxidative phosphorylation pathway. For each sample in [U-^13^C]glucose labeled NSCLC dataset (top) and RCC (bottom), true ranks of citrate m+2 were compared with predicted ranks from UnitedMet (left) and oxidative phosphorylation pathway scores calculated from gene expressions in the corresponding Hallmark gene set (right). **d)** UMAP plots of sample embedding matrix W (posterior means) learned by UnitedMet reveal integration of batches (top) and clustering across renal cell carcinoma subtypes in the TCGA-KIPAN batch (bottom). Each dot represents a patient sample. RCC and TCGA-KIPAN samples overlap in the latent space. **e)** UnitedMet captures histology-associated differences in metabolism across RCC subtypes. Differential abundance of imputed isotopologues across RCC subtypes in TCGA KIPAN were compared to ground-truth differences in the measured RCC cohort. Isotopologues in blue are consistently and significantly enriched (FDR-adjusted p < 0.1, Wilcoxon rank-sum test) in both measured and predicted cohorts. **f)** UnitedMet captures mutation-driven metabolic reprogramming in ChRCC. For each sample in the ChRCC cohort (n=61), predicted levels of lactate m+3/glucose m+6 (left) and citrate m+2/glucose m+6 (right) are shown. Error bars represent ± one standard deviation. X-axis is sorted by predicted abundances of corresponding isotopologues. Samples with complex 1 insertion or deletions are labeled red. Samples with complex 1 single nucleotide variations are labeled green. p values show the results of Wilcoxon rank sum test between complex 1 indel samples and the other samples.

Human kidney cancer arises in a variety of subtypes, including clear cell (ccRCC), papillary (pRCC), and chromophobe (chRCC), presenting with functionally distinct metabolic activity. To further benchmark the capacity of UnitedMet to impute isotopologue distributions, we assessed its capacity to capture histology-associated differences in metabolism across RCC subtypes. To do so, we applied UnitedMet, using multimodal RNA-seq/isotopologue data from Bezwada et al ^24^ as a reference dataset, and 1020 RCC tumor and adjacent normal samples from the TCGA Pan-kidney cohort (KIPAN) (encompassing ccRCC, pRCC, and chRCC) as a target dataset (**Figure S3B**). At the low-dimensional latent space learned by UnitedMet, we found that UnitedMet successfully embedded samples in both the reference and target datasets according to their subtype, despite missing measurements of isotopologues in the TCGA KIPAN (**Figure 3D**). Furthermore, imputed labeling patterns in the TCGA KIPAN dataset preserved ground-truth differences between both ChRCC/ccRCC samples (Spearman ρ = 0.85, p = 4.2 × 10^−15^) and pRCC/ccRCC samples (Spearman ρ = 0.79, p = 5.0 × 10^−12^) (**Figure 3E**). Consistent with prior findings ^24^, ccRCC samples demonstrated higher glycolytic labelings such as lactate m+3/glucose m+6, while ChRCC and pRCC samples displayed higher ratios of TCA cycle labelings such as citrate m+2/glucose m+6 and succinate m+2/glucose m+6 (**Figure 3E**). While ChRCC displays increased utilization of the TCA cycle, loss-of-function alterations to mitochondrial DNA (mtDNA)-encoded Complex I genes can result in loss of oxidative phosphorylation and metabolic reprogramming to glycolytic pathways ^25^. Consistent with these findings, we found ChRCC samples with Complex I alterations demonstrated a shift to an alternative glycolytic metabolic pathway with higher levels of lactate m+3/glucose m+6 (p = 0.02) and lower levels of citrate m+2/glucose m+6 (p = 0.04, Wilcoxon rank sum test, **Figure 3F**). This suggested that UnitedMet captured mutation-driven metabolic reprogramming in ChRCC, which further validated UnitedMet’s capability to generate biologically meaningful predictions.

In total, the analysis presented in **Figure 2** and **Figure 3** demonstrates that UnitedMet is capable of accurately imputing both metabolite levels as well as isotopologue distributions from RNA-seq data via joint, multimodal modeling with reference datasets.

### BAP1 mutations are associated with an oxidative metabolic phenotype in ccRCC

Although both oncogenes (such as *MYC* and *PIK3CA*) and tumor suppressor genes (*PTEN, VHL*) are well-recognized regulators of metabolism ^26,27^, the functional consequences of driver alterations on tumor metabolism *in vivo* is poorly studied ^28,29^. In fact, the lack of population-scale metabolomic profiling in contemporary cohorts of molecularly profiled tumors renders a direct evaluation of the association of either metabolite levels or metabolic flux with the presence of specific driver alterations infeasible. We reasoned that we could apply UnitedMet to impute both metabolite levels and isotope labeling patterns in richly profiled cohorts of tumors, such as those from the TCGA, to assess whether genomic alterations were associated with specific metabolite changes.

We focused our efforts on understanding the genome-metabolome covariation in ccRCC, for which we have several reference datasets with both transcriptomic and metabolomic/labeling data. The canonical founder mutation in ccRCC is the biallelic inactivation of the tumor suppressor gene *VHL* and the activation of a pseudohypoxic transcriptional and metabolic program. The subsequent evolution of ccRCC includes the acquisition of secondary driver mutations in genes (such as *PIK3CA, PTEN, MTOR*, and *BAP1*) whose functions are (at least in part) metabolic^2^. To understand the associations between genetic mutations and metabolic variations in ccRCC, we applied UnitedMet to large-scale multi-omics TCGA Kidney renal clear cell carcinoma (KIRC) cohort (n=606) which has paired RNA-seq and whole exome-sequencing (WES) data. Training the RNA-seq data from TCGA KIRC with 4 ccRCC reference datasets (CPTAC, CPTAC_val, RC18, RC20, n=341) containing paired RNA-seq and metabolomics data, UnitedMet predicted metabolite levels for TCGA KIRC samples(**Figure 4A**) Similarly, UnitedMet predicted isotopologue distributions for TCGA KIRC samples by training them with the [U-^13^C]glucose labeled reference dataset RCC (**Figure 4E**).

**Figure 4.**
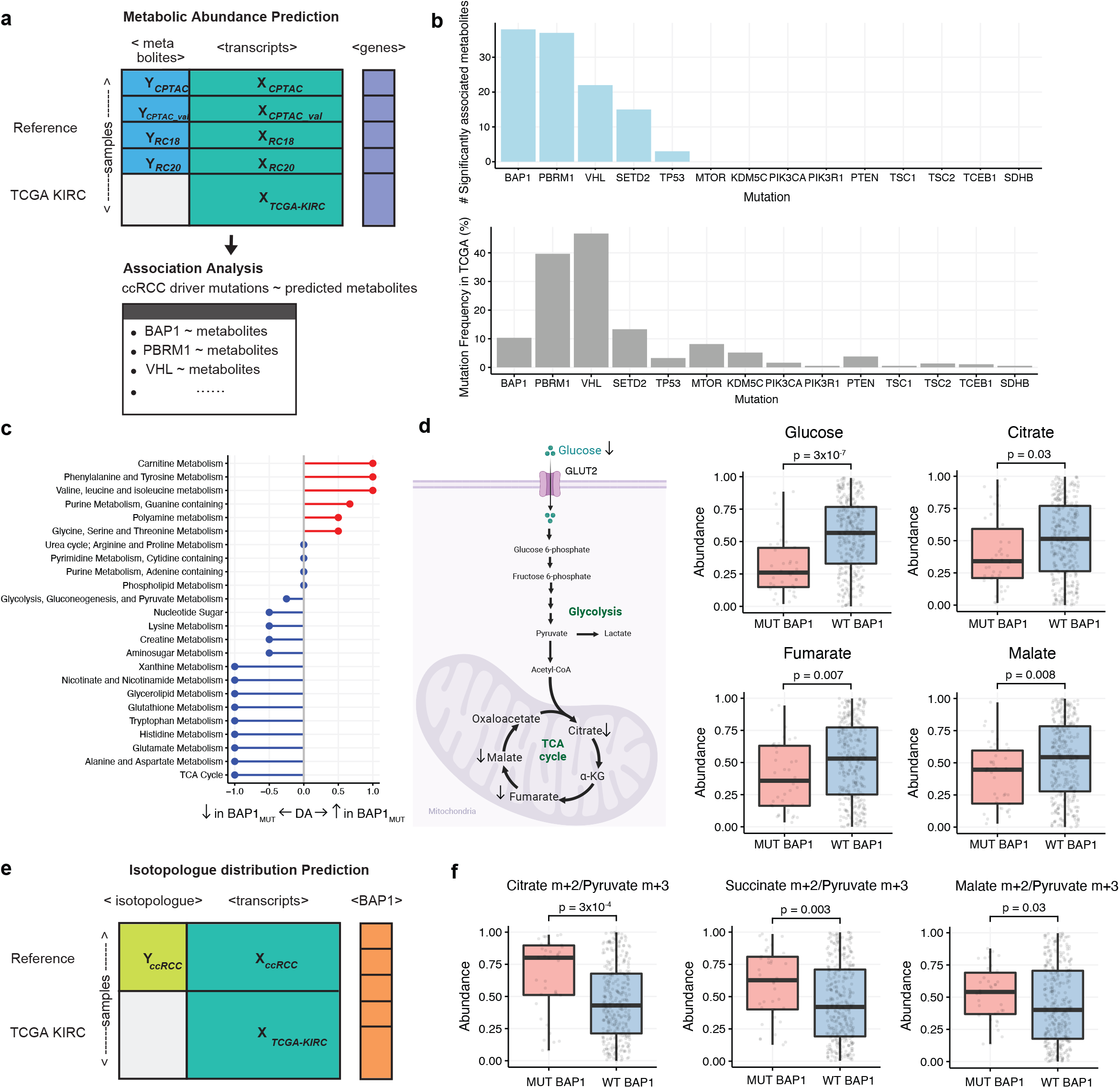
BAP1 mutations in ccRCC are associated with a unique metabolic phenotype. **a)** Schematic of the metabolite level prediction and downstream analysis for TCGA KIRC samples with UnitedMet. RNA-seq data (*X*_*TCGA*_) of the TCGA KIRC cohort (target dataset) are trained with 4 ccRCC reference datasets (CPTAC, CPTAC_val, RC18, RC20, n=341) containing paired RNA-seq and metabolomics data. *X*: RNA-seq data; *Y*: Metabolomics data. Predicted metabolite levels (*Y*_*TCGA*_) are leveraged for association analysis with ccRCC driver mutations. **b)** BAP1 mutation demonstrates the strongest association with a broad range of predicted metabolites. Distribution of the total number of significantly associated metabolites across 14 key driver mutations in ccRCC (top). X-axis is sorted by the number of significantly associated metabolites. Mutation frequency of 14 driver genes (bottom). X-axis is sorted by mutation frequency. **c)** Pathway-based analysis of predicted metabolic changes in BAP1 mutant v.s. BAP1 wildtype samples in TCGA KIRC cohort. **d)** Predicted metabolite level changes in BAP1 mutant v.s. BAP1 wildtype samples in TCGA KIRC cohort. Left: Diagram of glucose metabolism pathways: glycolysis and TCA cycle. Right: Boxplots comparing predicted unphosphorylated glucose, citrate, fumarate malate levels in MUT BAP1 v.s. WT BAP1 samples. p values are calculated by unpaired two tailed parametric t-tests. **e)** Schematic of the isotopologue distribution prediction for TCGA KIRC samples with UnitedMet. RNA-seq data (*X*_*TCGA*_) of the TCGA KIRC cohort (target dataset) are trained with ccRCC samples in the [U-^13^C]glucose labeled RCC dataset containing paired RNA-seq and isotope labeling data. *X*: RNA-seq data; *Y*: Isotopologue data. **f)** Predicted isotopologue changes in BAP1 mutant v.s. BAP1 wildtype samples in TCGA KIRC cohort. Boxplots comparing predicted citrate m+2/pyruvate m+3, succinate m+2/pyruvate m+3, malate m+2/pyruvate m+3 ratios in MUT BAP1 v.s. WT BAP1 samples. P values are calculated by unpaired two tailed parametric t-tests.

We first studied associations between the predicted metabolite abundances and genetic mutations in the TCGA KIRC cohort. For each reproducibly well-predicted metabolite, we compared the predicted abundances in mutant samples to wild-type samples among fourteen key driver mutations in ccRCC (*VHL, PBRM1, SETD2, BAP1, MTOR, KDM5C, PTEN, TP53, PIK3CA, TSC2, TCEB1, TSC1, PIK3R1, SDHB*), using a false detection rate (FDR)-corrected Wilcoxon test (**Figure 4B**). We identified significant higher/lower mutation-specific abundance of metabolites in *BAP1* (n=38 metabolites), *PBRM1* (n=37), *VHL* (n=22), *SETD2* (n=15), and *TP53* (n=3) mutations. *BAP1* mutation showed the strongest association with the largest variety of predicted metabolites despite a relatively low mutation rate (∼10%) in ccRCC patients (**Figure 4B**). Consistent with this observation, prior literature has shown that *BAP1* mutations play a role in several aspects of cellular metabolism including glucose metabolism ^30^. Mass spectrometry measurements demonstrated that germline *BAP1* mutations induced the Warburg effect in human fibroblasts, including depleted TCA cycle activity and increased aerobic glycolysis ^31^. Additionally, transcriptome analysis showed that *BAP1*-mutant ccRCC patient samples were enriched in glycolytic gene expression ^32^. To gather insight into the interplay between *BAP1* mutation and metabolite abundance, we performed a pathway-based differential abundance (DA) analysis of predicted metabolic changes in *BAP1* mutant and wild-type samples in TCGA KIRC. *BAP1* mutant samples showed significant depletion in the TCA cycle metabolism (DA score = −1), including drops in the levels of citrate (p = 0.03), fumarate (p = 0.007), and malate (p = 0.008)(**Figure 4C, Figure 4D**). *BAP1*-mutant samples also demonstrated lower levels of free, unphosphorylated glucose (p = 3 × 10^−7^), suggesting that these tumors may upregulate glucose uptake from the microenvironment (**Figure 4D**).

To more granularly understand the metabolic flux patterns associated with the above-described pool size changes, we leveraged imputed [U-^13^C]glucose-labeled isotopologue distribution data from TCGA KIRC. Relative to *BAP1-*wild-type tumors, *BAP1-*mutant tumors demonstrated increased levels of citrate m+2/pyruvate m+3 (p = 3 × 10^−4^), succinate m+2/pyruvate m+3 (p = 0.003), malate m+2/pyruvate m+3 (p = 0.03) (**Figure 4F**), indicating an elevated contribution of glucose to TCA cycle activity in *BAP1*-mutant ccRCC. These data indicate that pool size drops in TCA cycle metabolites are not caused by decreased entry of glucose into the TCA cycle. Instead, they suggest that *BAP1-*mutant tumors undergo reduced entry of other anapleurotic sources of TCA cycle intermediates, such as glutamate, or alternatively increase diversion of TCA cycle intermediates into alternate pathways, such as the utilization of acetyl-CoA for fatty acid synthesis. Such hypotheses are directly testable by analogous infusion experiments using, for example, labeled glutamine, and suggest that *BAP1* tumors may harbor metabolically distinct (and potentially therapeutically targetable) metabolic alterations.

### Shift to oxidative metabolism correlates with disease progression and poorer clinical outcome

Recent work has suggested that, although ccRCC tumors generally downregulate mitochondrial gene expression and limit entry of glucose-derived carbon into the TCA cycle relative to normal tissue, distant metastases in ccRCC upregulate oxidative phosphorylation and glucose entry into the TCA cycle ^24^. However, there is no large-scale data available on the metabolism of metastatic tumors. We reasoned that high-stage, aggressive ccRCC tumors, which ultimately seed distant metastases, should exhibit signatures of upregulation of oxidative glucose metabolism. To test this hypothesis, we again leveraged predicted isotopologue distribution data from TCGA KIRC and compared isotopologue levels of [U-^13^C]glucose-labeled TCA cycle intermediates normalized by pyruvate m+3 in ccRCC tumors from different pathological stages. Aggressive ccRCCs with higher stage demonstrated higher levels of citrate m+2/pyruvate m+3 (p = 3 × 10^−4^), succinate m+2/pyruvate m+3 (p = 7 × 10^−6^) and malate m+2/pyruvate m+3 (p = 2 × 10^−4^, Kruskal–Wallis test; **Figure 5A**), consistent with increased glucose-derived carbon entry into the TCA cycle. We then applied UnitedMet to predict isotopologue distribution data for 823 primary or metastatic tumor samples from a publicly available advanced ccRCC clinical trial (IMmotion151). We trained RNA-seq data from IMmotion151 with the ccRCC samples from the RCC reference dataset. Predicted isotopologue levels of [U-^13^C]glucose-labeled TCA cycle intermediates normalized by pyruvate m+3 were compared between primary and metastatic ccRCC tumors in IMmotion 151. Metastatic ccRCC tumor samples demonstrated higher levels of citrate m+2/pyruvate m+3 (p = 5 × 10^−13^), succinate m+2/pyruvate m+3 (p = 3 × 10^−10^) and malate m+2/pyruvate m+3 (p = 2 × 10^−9^, Wilcoxon rank sum test; **Figure 5B**). Together, these results indicate that, in ccRCC, increased TCA cycle activity is associated both with (1) high stage/disease progression and (2) the establishment of metastasis itself.

**Figure 5.**
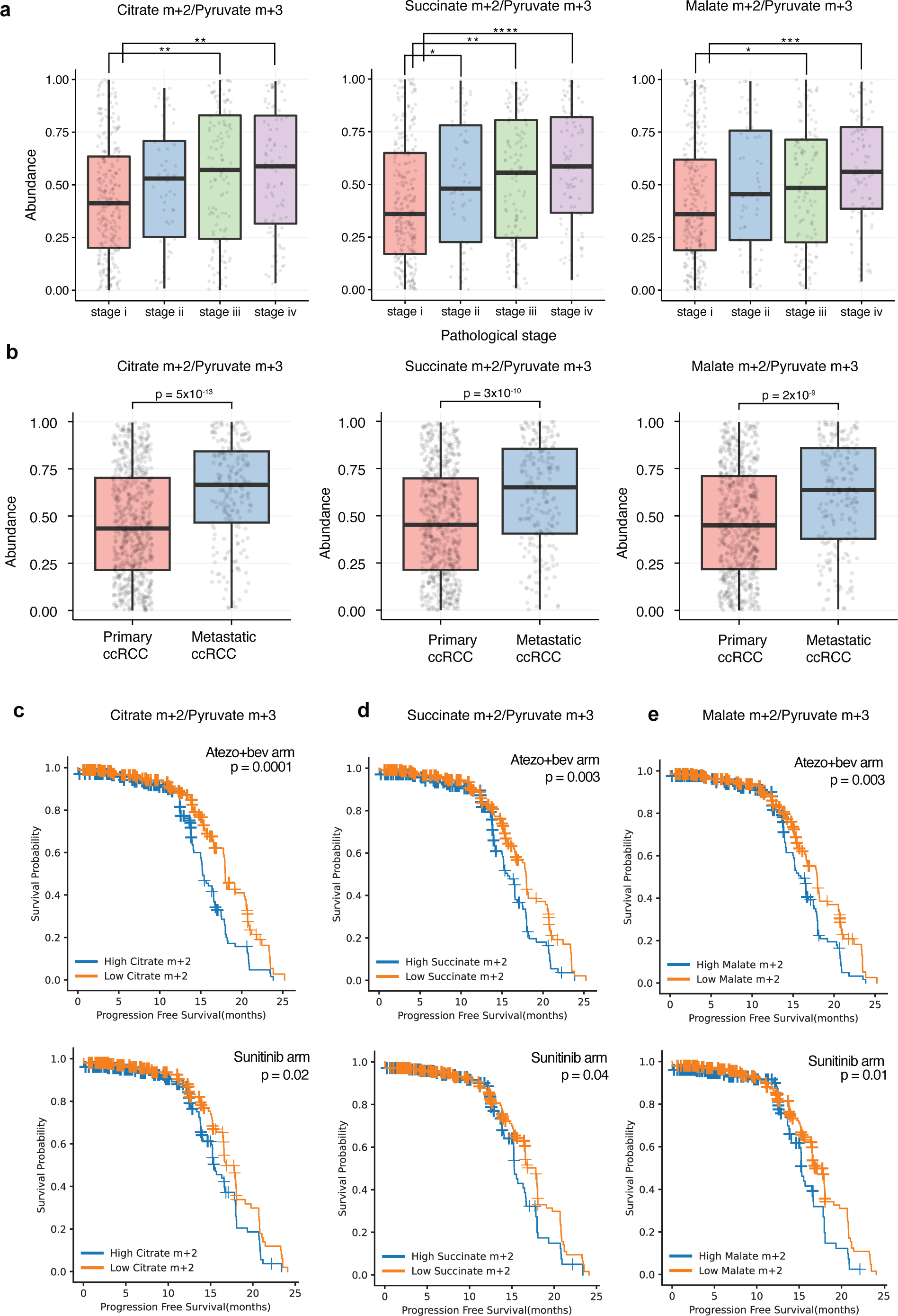
Shift to oxidative metabolism correlates with disease progression and poorer clinical outcome. **a)** Aggressive ccRCCs with higher stage demonstrate higher ratios of predicted citrate m+2/pyruvate m+3 (left), succinate m+2/pyruvate m+3 (middle) and malate m+2/pyruvate m+3 (right) in TCGA KIRC cohort. Significance between any two stages is assessed by pairwise t-test. P values are FDR adjusted. * *P* < 0.1, **** *P* <0.01, **** *P* < 0.001, **** *P* < 0.0001. **b)** Samples from metastatic sites in ccRCC patients show higher ratios (compared to samples from primary tumor sites) of predicted citrate m+2/pyruvate m+3 (left), succinate m+2/pyruvate m+3 (middle) and malate m+2/pyruvate m+3 (right) in IMmotion151 cohort. Significances are assessed by the Wilcoxon rank sum tests. **c)** Kaplan-Meier plot showing that ccRCC patients with a high level of citrate m+2/pyruvate m+3 (based on median level) had poorer PFS than patients with low level of citrate m+2/pyruvate m+3 in both atezo+bev arm (top) and sunitinib (bottom) arm. Significance is assessed by the log-rank test. **d)** Same as c) but for succinate m+2/pyruvate m+3. **e)** Same as c) but for malate m+2/pyruvate m+3.

We next interrogated if this oxidative metabolic phenotype may be linked to poor clinical outcomes. ccRCC patients in the IMmotion151 trial were treated with either atezolizumab plus bevacizumab (a combination of tyrosine kinase inhibitor and immunotherapy) or sunitinib (a tyrosine kinase inhibitor). We evaluated the association between isotopologue levels of TCA cycle intermediates and progression-free survival (PFS) by multivariate Cox proportional hazards models (evaluating different treatment arms separately). We observed that patients with high citrate m+2/pyruvate m+3 (p = 0.0001 in atezo+bev arm, p = 0.02 in sunitinib arm, **Figure 5C**), succinate m+2/pyruvate m+3 (p = 0.003 in atezo+bev arm, p = 0.04 in sunitinib arm, **Figure 5D**) and malate m+2/pyruvate m+3 (p = 0.003 in atezo+bev arm, p = 0.01 in sunitinib arm, **Figure 5E**) were all associated with poorer PFS in both arms. These data nominate oxidative metabolism of glucose as a potential druggable target to diminish cancer progression and metastasis in patients receiving both immunotherapy and anti-angiogenic agents in ccRCC.

## Discussion

This work presents a novel methodology for the joint, probabilistic modeling of multimodal metabolic data. In doing so, it addresses the numerous challenges associated with analysis of metabolomics data (including but not limited to semi-quantitative data and batch effects) and its joint modeling with transcriptomics data. After establishing that UnitedMet accurately imputes metabolite features in benchmark datasets, we applied UnitedMet to study the metabolic consequences of key driver mutations and the metabolic adaptations associated with aggressive disease and metastatic competency.

Several key limitations underlie UnitedMet and represent important challenges in development of next-generation methods for joint modeling of multimodal metabolic data. Among these, two are of significant near-term importance. First, while rank transformation has proven useful in both UnitedMet and MIRTH ^23^ for the comparison of semi-quantitative metabolite data produced in distinct batches, the process of rank transformation produces a loss of information, where large effect sizes (*i*.*e*. large fold changes between pairs of samples) in one metabolite feature can be equated to small effect sizes in another metabolite feature in the rank-transformed space. Second, UnitedMet requires at least one reference dataset of sufficient size in order to carry out imputation. For the majority of diseases, such a dataset does not exist ^14,33^. One potential avenue to overcoming this challenge is to train disease-agnostic models to impute metabolite features. This seems feasible for at least some metabolite features that demonstrate lineage-agnostic covariation with gene expression, such as *IDO1* and kynurenine ^14,22^.

UnitedMet harnesses the covariation between the transcriptome and metabolome to impute otherwise-unmeasured metabolite features. In doing so, it enables the inference of pool size and tracing patterns (and, by consequence, the evaluation of metabolite-centered hypotheses) in samples where metabolite profiling is difficult or otherwise infeasible. Several valuable use cases come to mind as natural applications of the UnitedMet framework where ancillary transcriptomic data is available. For instance, one may seek to infer metabolite levels in archival FFPE samples of inadequate quality for metabolite profiling, or in biopsy samples with an inadequate quantity of material for metabolite profiling. Separately, in isotope-tracing experiments where one is interested in more than one tracer (*e*.*g*. ^13^C-glucose and ^13^C-glutamine tracing, where infusion of both tracers in the same patient does not provide useful data), UnitedMet could be used to impute the outcome of the counterpart tracer as long a common data modality (*e*.*g*. RNA-seq) was collected. UnitedMet therefore democratizes metabolomics data for scientific discovery.

## Methods

### Data preprocessing

The input to UnitedMet consists of reference datasets with paired measurements of RNA counts and total ion counts of metabolites/isotopologues and a single-modality target dataset with RNA-seq data only (**Figure 1A**). We assume that there are *N* different reference datasets, each with a RNA-seq sample×gene matrix of raw counts 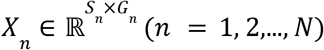 and a paired sample×metabolite or sample×isotopologue ion count matrix 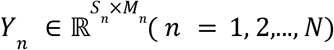. Let 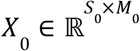 be the RNA-seq sample×gene matrix in a single-modality target dataset.

#### Normalization

We first normalized all input data with distinct techniques. We implemented total ion count (TIC) normalization to raw ion count matrices of metabolomics data (*Y*) and transcripts per million (TPM) normalization to raw count matrices of RNA-seq data (*X*). In metabolomics experiments, ion counts below a threshold were not detected by the mass spectrometry. This ended up with missing metabolite measurements in some samples. We treated these left-censored values as half of the minimum value across all metabolite measurements when calculating the TIC normalizer.

For sample×isotopologue ion count matrices (*Y*) of isotope labeling data, we first calculated the fractional labeling, namely the proportion of each isotopologue relative to the sum of all isotopologues in that metabolite. We then divided all fractions by the fraction of pyruvate m+3 or glucose m+6. Normalization by pyruvate m+3 allowed us to establish the labeling ratio of each isotopologue to pyruvate m+3, providing insights into the contribution of glucose-derived pyruvate to that specific isotopologue.The labeling ratio of citrate m+2 to pyruvate m+3, for instance, suggested the contribution of glucose through the pyruvate dehydrogenase (PDH) reaction. Normalization by glucose m+6 instead revealed the contribution of glucose carbon to other metabolites.

#### Rank-transformation

As metabolomics/isotope tracing data generated using mass spectrometry are reported as semi-quantitative relative abundances, we are only able to compare measurements of the same metabolite/isotopologue from different samples in the same dataset. To map metabolic relative abundances and gene expression levels into a shared measurement scale across all features and datasets, we rank the metabolite/isotopologue and gene expression levels across all the samples within each dataset. Ranks enable the comparison of features across datasets and transfer learning from RNA-seq modality to metabolic modality. Samples exhibiting the maximum level for a specific feature within the provided dataset are assigned the highest rank. Conversely, samples displaying the minimum level for the same feature are allocated the lowest rank. Left-censored samples are tied, sharing the last rank in the ranking hierarchy. While we use unnormalized rankings for modeling, we normalize ranks by their total number of samples *N* in downstream analyses, mapping them to a comparable scale of ranks [0, 1) in all datasets. For each feature *j*, the normalized rank of a measurement *f*_*ij*_ (*i* = 1, 2,…, *N*) in that dataset is defined by 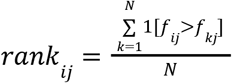.

#### Data aggregation

Rankings data of RNA-seq matrices *X*_*n*_ and metabolic matrices *Y*_*n*_ (*n* = 1, 2,…, *N*) in reference datasets are aggregated into a single data matrix *R* along with the rankings data of metabolic matrix *X*_0_ in the target dataset. While we take in the common genes shared across datasets to save computation costs, we aggregate metabolic modalities by embracing the union of relevant features. Namely, the aggregated matrix 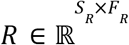, where 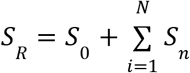 and *F*_*R*_ = (*G*_*R*_ + *M*_*R*_),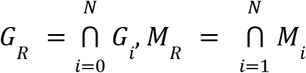. In the benchmarking test on four ccRCC datasets, R contains measurements of 1148 metabolites and 20171 genes for 341 samples.

### The UnitedMet model

UnitedMet is a probabilistic generative method that jointly models RNA-seq and metabolic data. UnitedMet assumes rankings in *R* are generated by a Plackett-Luce ranking distribution of a latent variable matrix *Z*, where *Z* = *W* ^*T*^*H* is the product of the latent sample embedding matrix 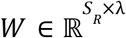 and the latent feature embedding matrix 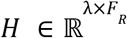. The hyperparameter λ is the number of embedding dimensions. We suppose all latent variables in both latent embedding matrices are generated by normal prior distributions:

*W*_*ik*_ ∼*Normal*(0, 1), *H*_*kj*_ ∼*Normal*(0, 1), where *W*_*ik*_ is the entry in the *i*^*th*^ sample and the *k*^*th*^ embedding column in embedding matrix W and *H*_*kj*_ is the entry in the *k*^*th*^ embedding row and the *j*^*th*^ feature in embedding matrix H.

#### Plackett-Luce ranking distribution

The Plackett-Luce distribution models a ranking of *N* items as an ordered series of choices. It begins by choosing the top-ranked item from the entire set of *N* options, followed by choosing the second-ranked item from the remaining options and so on ^34^. Given a set of N options {*Q*_1_, …, *Q*_*N*_}, the probability of selecting the *i*^*th*^ item *Q*_*i*_ is defined as 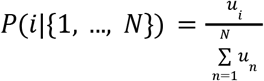 by the Luce Choice Axiom, where *u* represents the utility score of *Q*_*i*_. The probability of a full ordering {σ_1_, …, σ_*N*_}, where we assume 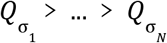, is then given by recursively applying the Plackett-Luce distribution: choose σ_1_ from {1, …, *N*}, choose σ_2_ from {1, …, *N*}\{σ_1_}, choose σ_3_ from 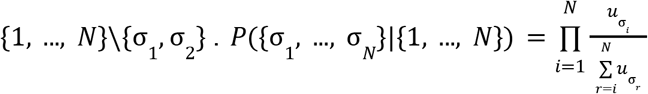. Given the latent variable matrix *Z* = *W*^*T*^ *H* in UnitedMet, we suppose the utility score of the item in the *i*^*th*^ sample and the *j*^*th*^ feature is defined as 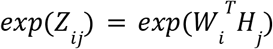. Extending this to censored rankings in UnitedMet, the likelihood of observing a censored ordering {σ_1_, σ_2_, …, σ_*K*_, {σ_*K*+1_,…, σ_*N*_}} in the *j*^*th*^ feature of a batch, is then defined by 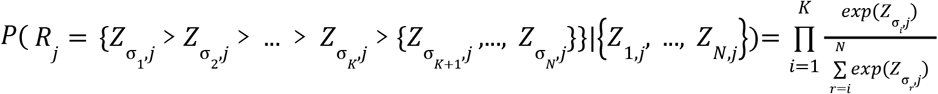. Detailed definitions of UnitedMet are described below.

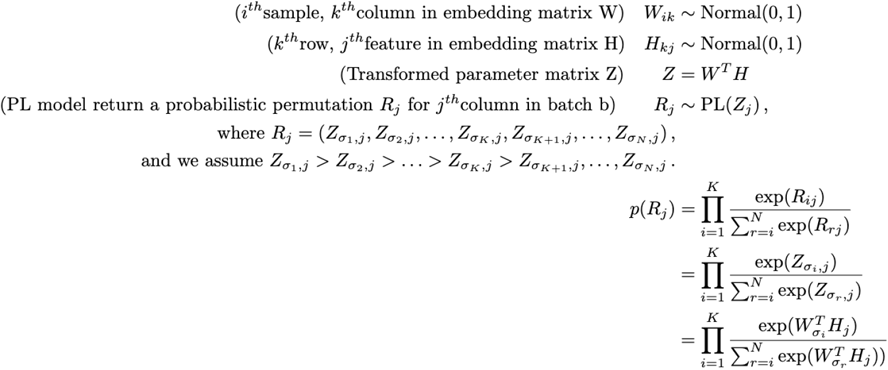

#### Cross-validation

To determine the optimal number of embedding dimensions (λ) of latent matrices *W* and *H*, we employ 10-fold cross-validation. The range of λ to be tested is contingent on the total number of samples *S*_*R*_. For instance, performance evaluation spans a λ range of [1,110] with a step of 10 in the benchmarking test on ccRCC datasets. For each batch, cross-validation features that are used to test model performance are selected separately. Only metabolic features (metabolites/isotopologues) that are measured in at least one other batch are included. These features are then randomly distributed into 10 folds. We treat one fold at a time as unmeasured and hold out the fold’s features in the corresponding batch. Masked features are then predicted by UnitedMet. In the end, we calculate the mean absolute error (MAE) between the true ranks of held-out features in the fold and their predicted ranks. The MAE scores across all folds are averaged to obtain a final performance score. We evaluate the MAE scores for all λ values, and the one resulting in the elbow of the MAE score curve is chosen as the optimal number of embedding dimensions for the factorization.

#### Inference

The likelihood is computed from observed rankings in both paired modalities of reference datasets, and only in the respective RNA-seq modality of the target dataset. We employ stochastic variational inference within the Pyro package for inference. Variational distributions are generated using the AutoNormal function. Optimization is executed through the Adam optimizer, with a default learning rate set to 0.001. Convergence is ascertained when the relative change in Evidence Lower Bound (ELBO) falls below 0.01.

#### Posterior prediction

UnitedMet estimates the joint posterior distribution of the latent embedding matrix *W* and *H*. For every latent variable in *W* and *H*, we draw 1000 samples from their estimated posterior distribution. Given posterior samples of the latent matrix *Z* (= *W*^*T*^ *H*), posterior rankings are then generated by the Plackette-Luce ranking distribution. To sample in a computation-efficient way, we implemented the Gumbel-Max trick, which generates ordered samples from the Plackette-Luce ranking distribution by sorting the perturbed log-probability through the addition of independent variables from the Gumbel distribution^35^. Let *G*_1_, …, *G*_*N*_ ∼ *Gumbel*(0), *iid*. Let *Z*_*j*_ be the *j*^*th*^ column of the latent matrix *Z*. Set perturbed log-probability *U*_*i*_ = *Z*_*i,j*_ + *G*_*i*_. The ordered indices of the *j*^*th*^ column returned by sorting the perturbed log-probabilities {*U*_1_, …, *U*_*N*_} are equivalent to the orderings generated by the Plackett-Luce model given probabilities(utility scores) {*Z*_1,*j*_, …, *Z*_*N,j*_}. Namely, if 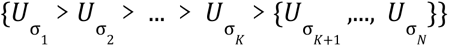, then we observe 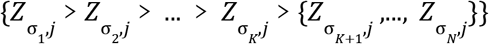. Estimates of the rankings can be found as the mean of the 1000 posterior draws, while the standard deviation of posterior samples represents a quantification of the prediction uncertainty.

### Benchmarking

#### Multivariable Lasso regression

We implemented multivariable Lasso regression on four ccRCC datasets according to Li et al^21^. In each dataset, metabolomics data were preprocessed by TIC normalization, while transcript levels are converted into TPM units. At each time in the benchmarking experiments, one ccRCC dataset was treated as the testing set while the other three were training sets. All RNA-seq data were scaled before training or testing. For every metabolite (y), we utilized gene expressions (x) to predict it in the training set. LassoCV in Python package scikit-learn was used to select the best penalizer alpha by 5-fold cross-validation. The maximum number of iterations fitting along the regularization path was set to default 1000. After selecting the best model for each metabolite, we assessed model accuracy by calculating spearman correlation coefficients between predicted metabolite levels and its ground truth.

#### MIRTH

MIRTH is a matrix factorization approach aimed at predicting the levels of unmeasured metabolites by collectively analyzing the co-variation of metabolites across multiple datasets^23^. We extended MIRTH to the cross-modality prediction problem. Metabolomics and RNA-seq data were preprossed in the same way mentioned above.

### Memorial Sloan Kettering Cancer Center (MSKCC) ccRCC datasets

We obtained two datasets RC18 (n=144) and RC20 (n=76), each with matched RNA-seq and mass spectrometry metabolomics measurements from fresh frozen high-quality tumor/adjacent normal specimens of ccRCC patients undergone partial or radical nephrectomies in MSKCC^22^. Samples were collected under the approval of Memorial Sloan Kettering Cancer Center’s institutional review board. The alignment of RNA-sequencing reads was performed using STAR 2-pass alignment against human genome assembly hg19. Metabolites were identified based on criterion according to Benedetti et al^14^. RC18 has measurements for 783 metabolites and 22937 genes. RC12 has measurements for 1012 metabolites and 22987 genes.

### CPTAC ccRCC datasets

Metabolite raw count matrices of CPTAC (n=50) and CPTAC_val (n=71) were downloaded from Li et al^1^. Transcriptomic and WES data were downloaded from Genomic Data Commons (GDC) at: https://portal.gdc.cancer.gov/projects/CPTAC-3 (Project: CPTAC-3, Primary Site: Kidney). CPTAC contained only ccRCC tumor samples, while CPTAC_val contained tumor and adjacent normal samples of ccRCC patients. Mass spectrometry peaks were quantified using Thermo Scientific Compound Discoverer® software to generate raw counts. HTSeq v0.11.2 was implemented to calculate the gene-level stranded read count. We then performed TIC normalization and TPM normalization on metabolite and gene expression count matrices respectively. CPTAC has measurements for 183 metabolites and 60483 genes. CPTAC_val has measurements for 130 metabolites and 60483 genes.

### Human RCC RNAseq + isotopic labeling data infused with [U-13C]glucose in vivo

Paired RNA-seq and isotopic labeling data from 76 primary tumor or adjacent normal kidney samples of RCC patients were downloaded from Bezwada et al^24^. The RCC dataset has measurements for 64 isotopologues and 12300 genes. Since small fluctuations of isotopologue levels that are not biologically interpretable can be quantified as signals in mass spectrometry, we set a criterion to filter out isotopologues whose average fraction over all samples are less than 10%. This ended up with a total of 23 isotopologues including biologically meaningful isotopologues like citrate m+2, malate m+2 etc.

### Human NSCLC cell line RNAseq + isotopic labeling data

We downloaded two human NSCLC cell line datasets with paired RNA-seq and isotopic labeling data from Chen et al^3^: NSCLC-G (n=85) and NSCLC-Q (n=85). 85 NSCLC cell lines were cultured with medium containing the isotopically enriched nutrient under identical conditions. The isotopic data in NSCLC-G was labeled by [U-13C]glucose, while the isotopic data in NSCLC-Q was labeled by [U-13C]glutamine. The NSCLC-G and NSCLC-Q dataset both have measurements for 78 isotopologues and 16383 genes.

### TCGA KIPAN datasets

We downloaded paired RNA-seq and WES data of 1020 RCC tumor and adjacent normal samples in TCGA KIPAN from the Genome Data Analysis Center (GDAC) in Broad Institute. 606 TCGA KIRC samples were included in TCGA KIPAN. mtDNA mutation calls using a Polymerase Chain Reaction (PCR)-based amplification approach for 61 ChRCC cases in TCGA KICH were downloaded from Davis et al^25^.

### Annotation of MAF files from WES data

We downloaded MAF files of WES data for CPTAC, CPTAC_val, TCGA KIPAN and TCGA KIRC from corresponding websites mentioned above. We annotated all molecular variations to 0 or 1 in a gene-wise way, where 0 represented wild-type or silent variations and 1 represented non-silent variations. Missense mutation, nonsense mutation, frame-shift deletion, splice site mutation, frame-shift insertion, in-frame deletion, splice-region variant, translation start site mutation, in-frame insertion and nonstop mutation were considered as non-silent molecular variations. Silent mutations, intron mutation, 3’ UTR mutation, and 5’ UTR mutation were considered as silent variations, because they were not able to change gene functions.

### Differential abundance score

The Differential Abundance (DA) score assesses the distinct regulation of a metabolic pathway between two groups. Calculated through a Wilcoxon rank sum test applied to all pathway metabolites, the score undergoes P-value correction using the Benjamini-Hochberg method (FDR-corrected p-value < 0.05). For each pathway, the DA score is derived as follows: (#significantly enriched metabolites - #significantly depleted metabolites) / #total metabolites. Scoring is exclusively applied to pathways exhibiting three or more significantly altered metabolites.

### Survival analysis

We collected RNA-seq data and patient-level clinical information from IMmotion151^36,37^ (N=823), a published trial exploring immunotherapeutic versus systemic agents in advanced ccRCC. To account for diverse drug effects in clinical trials, we conducted separate statistical analyses for the immunotherapy arm (Atezolizumab + Bevacizumab) and the sunitinib arm. The survival regression analysis was performed using the Python package lifelines.

### Statistical analysis

Statistical analyses were conducted using either R or Python. Differential distribution comparisons were implemented with Wilcoxon rank sum test or t-test. All statistical tests were two-sided by default, unless specified otherwise, with p-values corrected using the Benjamini-Hochberg method^38^.

## Data Availability

All data needed to evaluate the conclusions in the paper are present in the paper and/or available online at 10.5281/zenodo.11286535. UnitedMet is open-source and online at: https://github.com/reznik-lab/UnitedMet.

https://doi.org/10.5281/zenodo.11286535

https://github.com/reznik-lab/UnitedMet

## Supplemental Figures

**Supplemental Figure 1. UnitedMet outperforms Lasso and MIRTH at predicting metabolite levels from RNA-seq data. a)** UnitedMet’s hyperparameter λ, the number of embedding dimensions, is determined by 10-fold cross-validation in the benchmarking experiments of 4 ccRCC datasets. Average mean absolute error between predicted ranks and true ranks across 10 folds changes with different numbers of embedding dimensions. Optimal dimension is picked by the knee point of the curve. Performance evaluation spans a λ range of [1,351] with a step of 10. **b)** The imputation performance for each dataset is assessed by the number of well-predicted metabolites. Metabolites with predicted ranks that show significant positive correlation (FDR-adjusted p < 0.05 and spearman rho > 0) are defined as well-predicted. **c)** Performance to impute 50% held-out metabolites from the remaining 50% measured metabolites and RNA-seq data. Performance evaluated by the spearman rho among all predicted metabolites. Extended UnitedMet with a weighted loss function is used. d) Performance to impute 50% held-out metabolites from the remaining 50% measured metabolites and RNA-seq data. Performance evaluated by the number of well-predicted metabolites. Extended UnitedMet with a weighted loss function is used.

**Supplemental Figure 2. UnitedMet’s metabolite level predictions are consistent across 4 ccRCC datasets. a)** Correlation plots of metabolite-level prediction performances, characterized by their spearman correlation between true ranks and predicted ranks, in all pairwise comparisons of 4 ccRCC datasets. Each dot represents a metabolite. **b)** Prediction uncertainty is negatively correlated with the prediction accuracy. Prediction accuracy is estimated by quantifying the standard error of 1000 posterior draws of metabolite levels. For each metabolite, average standard error across all samples is associated with its prediction accuracy, characterized by spearman rho values between true ranks and predicted ranks. Each dot, colored by the proportion of censored measurements, represents a metabolite.

**Supplemental Figure 3. UnitedMet’s performance of predicting isotopologues from RNA-seq data. a)** UnitedMet’s hyperparameter λ, the number of embedding dimensions, is determined by 10-fold cross-validation in the benchmarking experiments of 3 isotope labeling datasets respectively. Performance evaluation spans a λ range of [1,81] with a step of 10. Average mean absolute error between predicted ranks and true ranks across 10 folds changes with different numbers of embedding dimensions. Optimal dimension is picked by the knee point of the curve. **b)** Schematic of the isotopologue distribution prediction for TCGA KIPAN samples with UnitedMet. RNA-seq data (*X*_*TCGA*−*KIPAN*_) of the TCGA KIPAN cohort (target dataset) are trained the [U-^13^C]glucose labeled RCC dataset containing paired RNA-seq and isotope labeling data. *X*: RNA-seq data; *Y*: Isotopologue data. Both target and reference datasets contain different subtypes of RCC. **c)** True ranks of Lactate m+3 were well-predicted by UnitedMet but not by gene expression signature of Hallmark glycolysis pathway. For each sample in [U-^13^C]glucose labeled RCC, true ranks of lactate m+3 were compared with predicted ranks from UnitedMet (left) and glycolysis pathway scores calculated from gene expressions in the corresponding Hallmark gene set (right).

## Acknowledgements

We thank Ralph Deberardinis, Brandon Faubert, Divya Bezwada for thoughtful feedback. Graphics were created with www.BioRender.com.

