## Supplementary figures and images for "UnitedMet harnesses RNA-metabolite covariation to impute metabolite levels in clinical samples"

### Supplemental figure 1

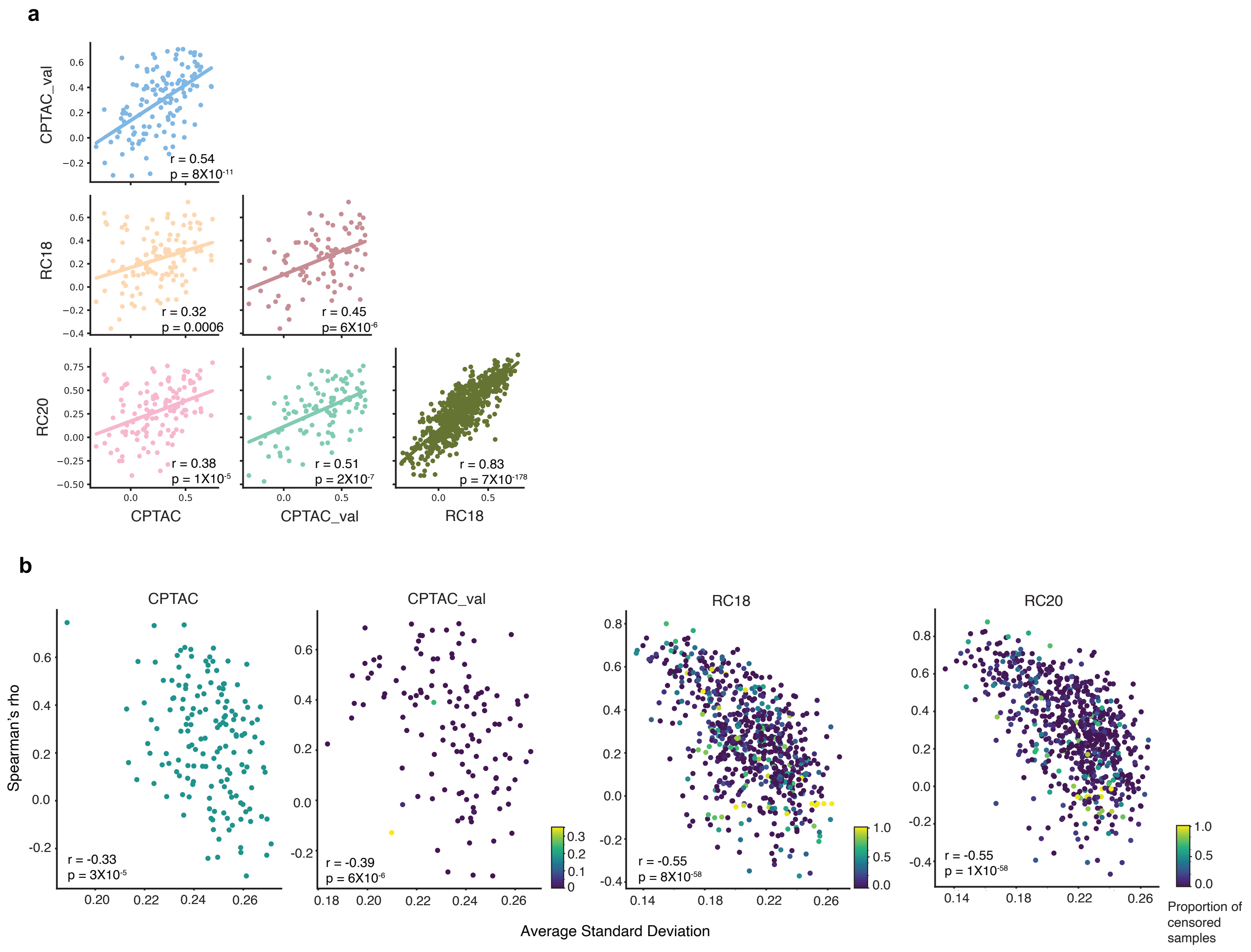

### Supplemental figure 2

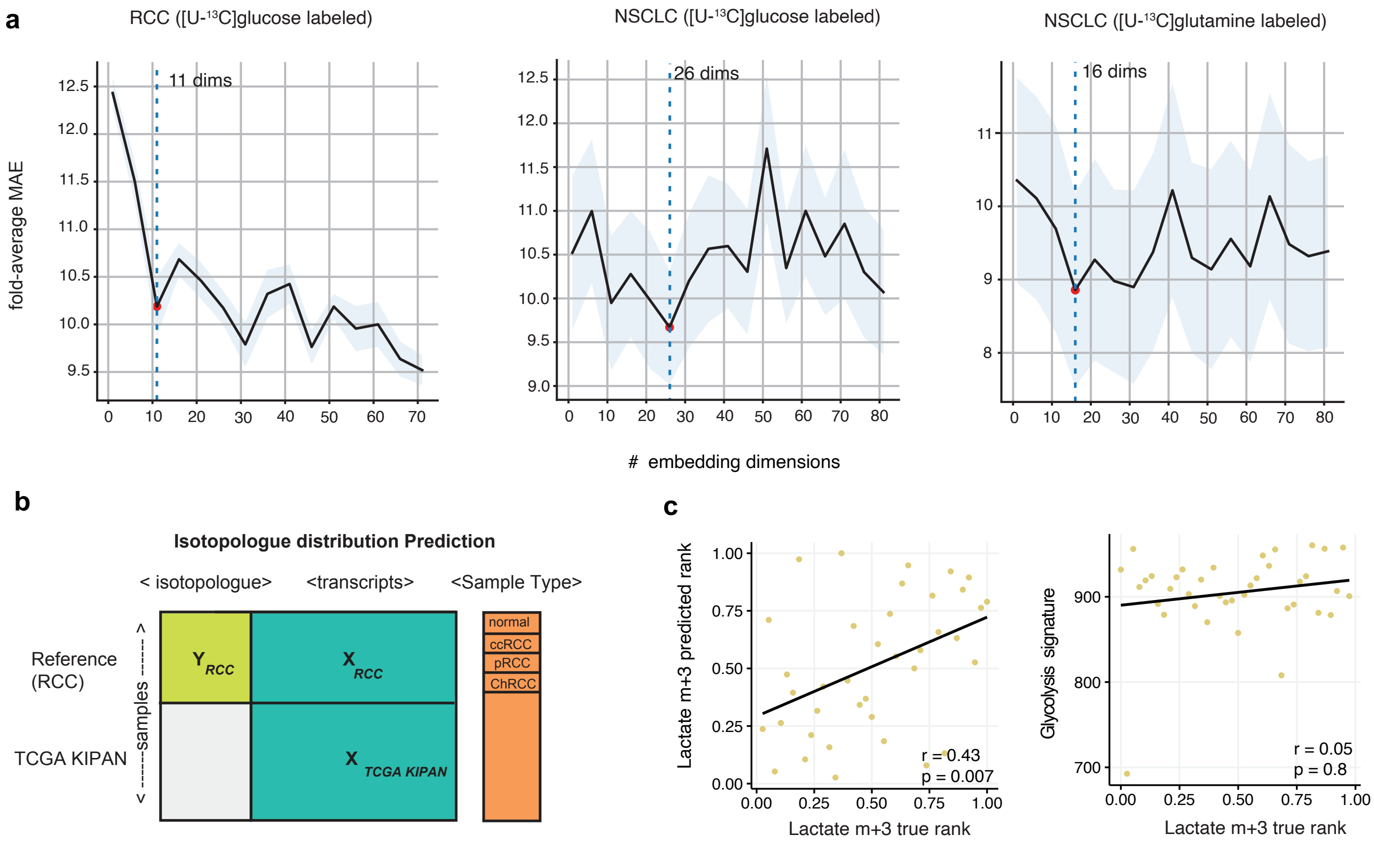

### Supplemental figure 3

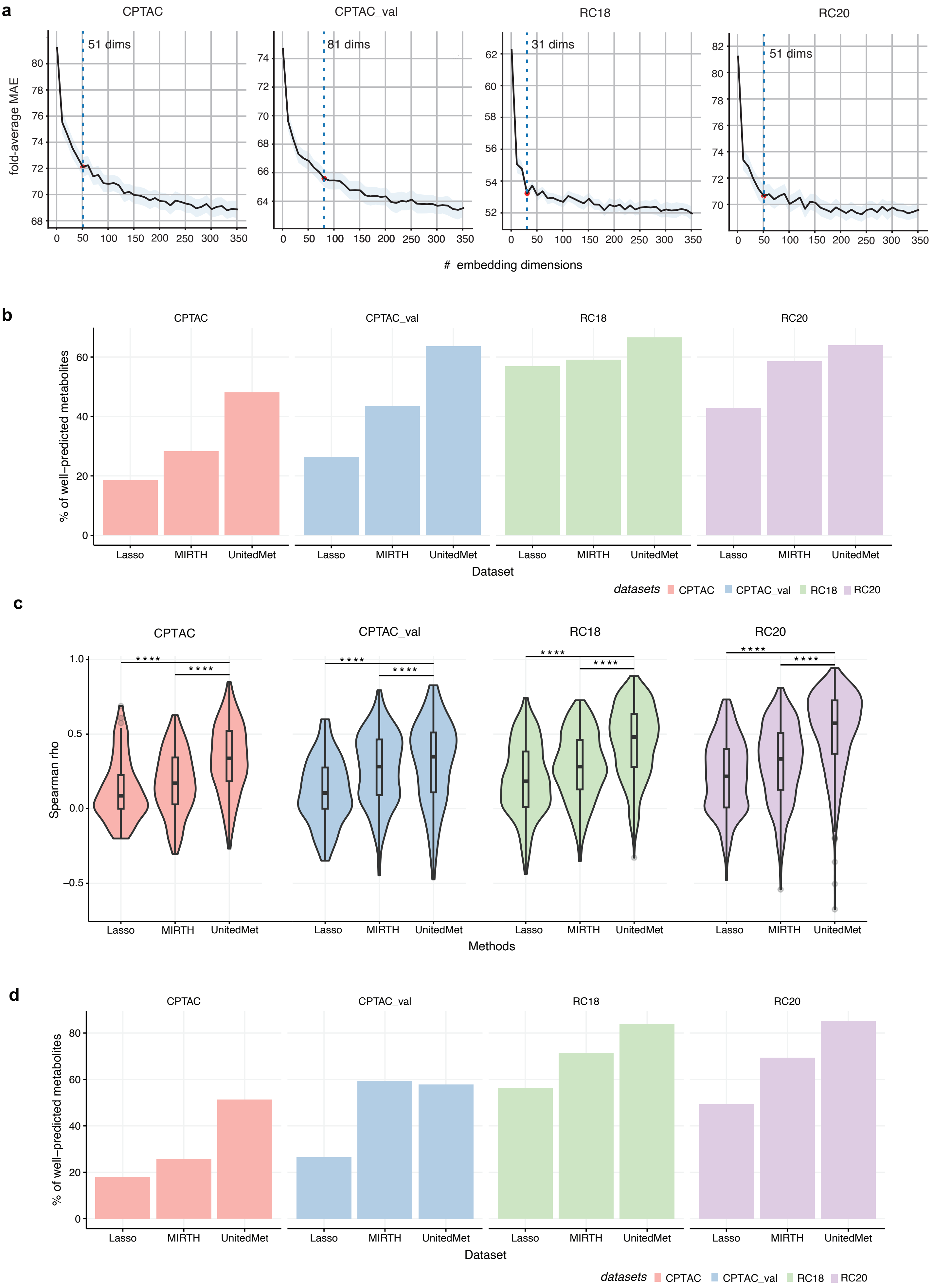
